# Shared genetic risk factors and their implications for treatment of IPF and systemic hypertension

**DOI:** 10.1101/2023.06.02.23290865

**Authors:** Gina Parcesepe, Richard J Allen, Beatriz Guillen-Guio, Samuel Moss, R Gisli Jenkins, Louise V Wain on behalf of the DEMISTIFI consortium

**Affiliations:** Department of Population Health Sciences, University of Leicester, Leicester, UK; NIHR Leicester Biomedical Research Centre, Leicester, UK; Margaret Turner Warwick Centre for Fibrosing Lung Disease, National Heart and Lung Institute, Imperial College London, London, UK

## Abstract

More than 50% of individuals with idiopathic pulmonary fibrosis (IPF) have co-morbid hypertension. High blood pressure can lead to organ fibrosis or can be a consequence of artery stiffening due to fibrosis. Studies have implicated common processes, such as TGF-β signalling, in both traits’ regulation. Our goal is to identify shared genetic risk factors for IPF and hypertension.

We analysed the genome-wide genetic correlation using LD Score Regression and the largest available genome-wide association studies of clinically defined IPF, and systolic and diastolic blood pressure (SBP and DBP, respectively). We then conducted a genome-wide colocalisation analysis to identify regions with a shared signal at P<10^−5^ between IPF and either SBP or DBP.

There was no genome-wide correlation between IPF and SBP (correlation (95% CI) -0.077(−0.142, -0.011), P=0.022) or DBP (correlation (95% CI) -0.027(−0.093, 0.039), P=0.427). The genome-wide colocalisation identified 8 shared signals, 3 (near *MAD1L1, GOLPH3L/HORMAD1*, and at 17q21.31) had the same direction of effect on risk of IPF and hypertension and 5 (near *TERC, OBFC1, DEPTOR*, and at 7q22.1 and 6p21.2) had opposite effects.

These findings support that there may be shared fibrotic mechanisms between IPF and hypertension. The opposite effects of variants at specific loci highlight the need for caution when considering therapeutic targeting of these shared pathways for either disease.

## Background

Idiopathic Pulmonary Fibrosis (IPF) is the most prevalent progressive idiopathic interstitial pneumonia (IIP) with a poor prognosis (median survival of 2.5-3.5 years from diagnosis and typically presents in older individuals (mean age=74). Systemic hypertension (high blood pressure) is highly prevalent and the leading contributor to all-cause death and disability worldwide. Hypertension is a risk factor for common comorbidities of IPF including ischaemic heart disease and more than 50% of individuals with IPF have systemic hypertension (defined as ≥130 mm Hg systolic blood pressure (SBP) and/or ≥80 mm Hg diastolic blood pressure (DBP) [1]). High blood pressure can lead to organ fibrosis or can be a consequence of artery stiffening due to fibrosis [2], and studies have implicated common pathways in both IPF and blood pressure regulation, such as TGF-β signaling and the renin-angiotensin system (RAS) [3]. We hypothesised that there could be shared genetic risk factors for both traits. Understanding the pleiotropic effects of the genes involved could have benefits and implications for drug repurposing and development in treating both IPF itself, and comorbid conditions.

## Methods

We utilised the largest available genome-wide association studies (GWAS) of clinically defined IPF [4], and SBP and DBP [5], to examine both genome-wide and locus-specific overlap. LD score regression (LDSC) [6] was used to conduct genome-wide correlation. Genome-wide colocalisation was applied using the coloc R package [7] to identify regions with a shared signal at P<10^−5^ between IPF and either SBP or DBP; this was performed by identifying signals at P<10^−5^ in both datasets, and then performing colocalisation. We report signals with a posterior probability of >80% for a shared causal variant (coloc H4) as significant. Signals were mapped to gene either using prior published evidence or using Open Targets Genetics (OTG). OTG utilises distance from gene, eQTL, splice QTL and functional annotation to suggest the most likely variant-to-gene mapping.

## Results

The IPF GWAS comprised 4,125 IPF cases and 20,464 controls. The SBP and DBP GWAS comprised 757,601 general population participants. There was no genome-wide correlation between IPF and SBP (correlation (95% CI) -0.077(−0.142, -0.011), P=0.022) or DBP (correlation (95% CI) -0.027(−0.093, 0.039), P=0.427). The genome-wide colocalisation analysis identified 18 regions of overlap of which 7 had a posterior probability of a shared causal variant >80% for IPF and blood pressure (Table 1). In addition, there was a shared signal at 17q21.31 for which colocalisation could not be reliably performed due to extended linkage disequilibrium across the region. However, the 17q21.31 signals for both IPF and blood pressure tagged an ancestral 1.5Mb genomic inversion [8] and so this was considered as a shared signal. Of the 8 shared signals, 3 (near *MAD1L1, GOLPH3L/HORMAD1*, and at 17q21.31) had the same direction of effect on risk of IPF and raised blood pressure (hypertension) and 5 (near *TERC, OBFC1, DEPTOR*, and at 7q22.1 and 6p21.2) had opposite effects with the allele associated with increased risk of IPF being associated with decreased blood pressure (and vice versa for the other allele) (Table 1).

**Table 1:** Colocalisation analyses results, stating the posterior probability that the traits are likely to share a causal variant.

| RSID | CHR | POS | Coded | Other | Coded allele frequency | IPF beta (se) | IPF Pvalue | SBP beta (se) | SBP Pvalue | H4 SBP (%) | DBP beta (se) | DBP Pvalue | H4 DBP (%) | Gene (source) |
| --- | --- | --- | --- | --- | --- | --- | --- | --- | --- | --- | --- | --- | --- | --- |
| rs3738485 | 1 | 150552392 | G | C | 0.45 | 0.134 (0.027) | 9.35x10<sup>-7</sup> | 0.164 (0.030) | 6.92x10<sup>-8</sup> | 90.6 | 0.018 (0.017) | 2.95x10<sup>-1</sup> | 75.9 | *GOLPH3L/HORMAD1* (OTG) |
| rs12696304 | 3 | 169481271 | G | C | 0.28 | 0.256 (0.030) | 8.45x10<sup>-18</sup> | -0.281 (0.035) | 4.23x10<sup>-16</sup> | 98.9 | -0.088 (0.020) | 9.94x10<sup>-6</sup> | 48 | *TERC* [4] |
| rs1214759 | 6 | 43352980 | G | A | 0.32 | 0.165 (0.029) | 1.71x10<sup>-8</sup> | -0.333 (0.032) | 4.79x10<sup>-25</sup> | 99.4 | -0.135 (0.019) | 2.51x10<sup>-13</sup> | 98.6 | 6p21.2 [4] |
| rs12699415 | 7 | 1909479 | A | G | 0.42 | 0.236 (0.027) | 7.85x10<sup>-18</sup> | 0.228 (0.031) | 1.82x10<sup>-13</sup> | 84.9 | 0.124 (0.018) | 3.30x10<sup>-12</sup> | 75.3 | *MAD1L1* [4] |
| rs2897075 | 7 | 99630342 | T | C | 0.38 | 0.263 (0.028) | 1.77x10<sup>-21</sup> | -0.156 (0.031) | 6.49x10<sup>-7</sup> | 99 | -0.101 (0.018) | 1.96x10<sup>-8</sup> | <1.0 | 7q22.1 [4] |
| rs28513081 | 8 | 120934126 | G | A | 0.43 | -0.180 (0.030) | 1.22x10<sup>-9</sup> | na | na | na | 0.084 (0.018) | 1.58x10<sup>-6</sup> | 89.6 | *DEPTOR* [4] |
| rs7100920 | 10 | 105640978 | T | C | 0.49 | 0.17 (0.027) | 1.67x10<sup>-10</sup> | -0.204 (0.030) | 1.62x10<sup>-11</sup> | 92.4 | -0.079 (0.017) | 5.43x10<sup>-6</sup> | 18.7 | *OBFC1* [4] |
| rs2077551 | 17 | 44214888 | C | T | 0.19 | -0.351 (0.038) | 1.92x10<sup>-20</sup> | -0.271 (0.042) | 8.75x10<sup>-11</sup> | na | -0.146 (0.024) | 1.24x10<sup>-9</sup> | na | 17q21.31 [4] |

RSID=reference SNP cluster ID, CHR=chromosome, POS=position (Build GRCh37), Coded=coded allele, Other=other allele, Coded allele frequency=coded allele frequency in IPF dataset, se=standard error, H4 is the posterior probability that the trait is likely to share a causal variant with IPF (probability >80% for a shared causal variant highlighted). Source is whether the gene information came from variant-to-gene mapping analyses from a prior paper [4] or Open Targets Genetics (OTG). OTG utilises distance from gene, eQTL, splice QTL and functional annotation to suggest the most likely variant-to-gene mapping.
na=not applicable. DEPTOR was not in the genome-wide colocalisation for SBP (not present at $P < 10^{-5}$ in dataset). Colocalisation was not performed for 17q21.31 due to extended linkage disequilibrium across the region.

## Discussion

Our study identified shared genetic associations between IPF and blood pressure traits with genetic variants at some loci increasing the risk of both IPF and hypertension, whilst at other loci, the same genetic variants increased risk of one trait whilst being protective against the other.

The DEP Domain-Containing MTOR-Interacting Protein (*DEPTOR*) encodes an mTOR inhibitor and decreased *DEPTOR* gene expression was associated with increased risk of IPF [9]. Targeting the mTOR pathway has been highlighted as a promising new therapeutic avenue for IPF [10]. Our findings suggest that intervening on this pathway may have potential adverse effects on blood pressure.

There was a significant difference in sample size for the GWAS used in our analyses and this will have affected the statistical power to detect signals in the smaller IPF dataset. Our genome-wide approach that utilised a more lenient threshold than the commonly used genome-wide threshold of P<5x10^−8^ will have partly mitigated this, but it is likely that more shared signals will be discovered as sample sizes increase. We only report signals with a single shared causal variant for IPF and blood pressure; it is plausible that nearby independent signals could exert their effect through the same nearby gene. It is likely that genetic effects may be tissue-specific and that different regulatory pathways may be involved in the expression of the same gene in different tissues.

Our findings provide support that drugs targeting shared mechanisms may have both anti-fibrotic and blood pressure lowering effects. However, the observation of opposite directions of effect for some genes suggests that blood pressure monitoring may be warranted in clinical trials for new IPF drugs that target genes that share genetic associations with blood pressure.

## Data Availability

All data produced in the present study are available upon reasonable request to the authors

## Data

IPF GWAS data are available from: https://github.com/genomicsITER/PFgenetics.

Blood pressure GWAS data are available from: https://www.ebi.ac.uk/gwas/publications/30224653

